# Oligogenic risk model for Gilles de la Tourette syndrome reveals a genetic continuum of tic disorders

**DOI:** 10.1101/2021.12.09.21266782

**Authors:** Malgorzata Borczyk, Jakub P Fichna, Marcin Piechota, Sławomir Gołda, Mateusz Zięba, Dzesika Hoinkis, Paweł Cięszczyk, Michal Korostyński, Piotr Janik, Cezary Żekanowski

## Abstract

Gilles de la Tourette syndrome (GTS) and other Tic Disorders (TDs) have a substantial genetic component with their heritability estimated at between 60 and 80%. Here we propose an oligogenic risk model of TDs using whole-genome sequencing (WGS) data from a group of Polish GTS patients, their families, and control samples (n = 278). The model is based on the overrepresentation of coding and non-coding genetic variants in and in the vicinity of genes selected from a set of 84 genes previously indicated as putatively associated with GTS. In the discovery phase, based on a variant burden test between unrelated GTS cases (n = 37) and a database of local allele frequencies 10 genes were selected for the model (*CHADL*, *DRD2*, *MAOA*, *PCDH10*, *HTR2A*, *SLITRK5*, *SORCS3*, *KCNQ5*, *CDH9,* and *CHD8*). Variants in these genes (n = 7654) with a median minor allele frequency in the non-Finnish European population of 0.02 were integrated into an additive classifier. This risk model was then applied to healthy and GTS-affected individuals from 23 families and 100 unrelated healthy samples from the Polish population (AUC-ROC=0.62, p=0.02). Application of the oligogenic model to a group of patients with other tic disorders revealed a continuous increase of the oligogenic score with healthy individuals with the lowest mean, then patients with other tic disorders, then GTS patients, and finally with severe GTS cases with the highest oligogenic score. Results were also overlapped with Psychiatric Genomics Consortium (PGC) GWAS data and we found no significant overlap between the common variant signal and our oligogenic model (p=0.21). Therefore obtained results were compared with the polygenic risk score built from the PGC GWAS data, which revealed a significant contribution of common variant background in severe GTS cases. Overall, we leveraged WGS data to construct a GTS/TDs risk model based on variants that may cooperatively contribute to the etiology of these disorders. This study provides evidence that typical and severe adult GTS as well as other tic disorders may exist on a single spectrum in terms of their genetic background.

## Introduction

Gilles de la Tourette syndrome (GTS) is a neurodevelopmental disorder characterized by multiple motor and at least one vocal and/or phonic tic which persists for longer than 12 months. The clinical phenotype of GTS belongs to the spectrum of tic disorders (TDs), a broad diagnostic category that includes: Chronic Motor Tic Disorder, Chronic Vocal Tic Disorder, Provisional Tic Disorder, Other Specified Tic Disorder and Unspecified Tic Disorder (Selvini, Cavanna, and Cavanna 2019). In about 90% of cases, GTS is accompanied by comorbid psychiatric disorders, including obsessive-compulsive disorder (OCD), attention-deficit and hyperactivity disorder (ADHD), autism spectrum disorder (ASD), affective disorders, anxiety disorders, impulse control disorders, and personality disorders (Robertson 2000). ADHD is associated with ca. 46% of GTS cases, and OCD with ca. 43% of cases (Hirschtritt et al. 2015). GTS and other TDs typically begin in childhood (average age at onset 4-6 years) or, less commonly, in adolescence. Most juvenile cases improve during adolescence and the prevalence of GTS in adulthood is about 20 times lower than in childhood (Bloch and Leckman 2009; Knight et al. 2012). Males are more commonly affected, with a male-to-female ratio of 3-4 to 1 (Freeman et al. 2000). The GTS prevalence in the general pediatric population ranges from 0.3% to 1% (Scharf et al. 2015). Other, non-GTS TDs, including isolated tics, are more common than GTS and affect up to 5% of the general population, however, this estimation varies depending on the methodology used (Robertson 2000). GTS and other TDs have a substantial genetic component, but the development of the clinical phenotype is complex and may be influenced by environmental, prenatal, and perinatal factors, hormonal disturbances, as well as psychosocial stressors (Gloor and Walitza 2016; Yu et al. 2019). The estimated heritability of GTS and other TDs based on twin, family, and population studies falls between 60 and 77% (Price et al. 1985; Yu et al. 2019).

Accordingly, candidate gene approach, linkage studies, and structural variant studies have shown that the genetic basis of GTS is heterogeneous and at least several genes have been implicated in the etiology of the disease (Pagliaroli et al. 2016; Qi et al. 2019). These genes are related to various processes and neurochemical pathways including dopaminergic, serotonergic, and histaminergic signaling (*DRD2, DRD4, SERT, HDC, DAT1, 5-HTTLPR*), as well as synapse development, remodeling and functioning (*SLITRK1, NLGN4, NRXN1*), differentiation of axons, cell adhesion (*CNTNAP2*), and mitochondrial activity (*IMMP2L*) (See also: Table 2) (Georgitsi et al. 2016; Qi et al. 2019). Still, the exact involvement of these genes in GTS etiology remains unknown, and the Human Phenotype Ontology database lists only two genes *HDC* and *SLITRK1*, as associated with GTS (Köhler et al. 2021). Recently, epigenetic mechanisms involving DNA methylation, histone acetylation, and gene regulation by non-coding RNAs have been proposed to mediate the impact of environmental factors on the genetic background of GTS (Qi et al. 2019).

It is now evident that no single gene is responsible for a large fraction of GTS cases, albeit rare variants with large effects have been considered causative in single GTS families (Baldan et al. 2014; Yuan et al. 2020). Genome-wide association studies (GWAS) of common neuropsychiatric disorders show that GTS is genetically correlated with OCD, ADHD, and major depressive disorder (MDD), diseases known to have an overlapping and highly polygenic background (Abdulkadir et al. 2018; Lee et al. 2019). The most recent GWAS of GTS included 6,133 GTS individuals and 13,565 ancestry-matched controls and after integrating genotyping data with transcriptomic and chromatin organization indicated *NR2F1* and *RBMS1* gene loci. GWAS-based polygenic risk scores (PRS) of GTS and other TDs calculated to date explain up to 3.3% of the variance in tic presence (Abdulkadir et al. 2019; Tsetsos, Topaloudi, et al. 2021; Yu et al. 2015). One of the hypotheses of the genetic background of GTS and non-GTS TDs states that these diseases are manifestations of varying severity on the same polygenic spectrum (Robertson et al. 2017).

Common variants with low impact identified by GWAS may contribute to the disease in a polygenic manner, while rare deleterious variants could contribute to GTS in a more family-specific manner. Here propose that a substantial part of GTS cases could be explained by an oligogenic model which assumes a compound effect of multiple low- and medium-impact variants with varied population allele frequencies. This hypothesis is supported by exomic data showing that *de novo* damaging variants in approximately 400 genes contribute to GTS risk in 12% of clinical cases (Willsey et al. 2017). GWAS results suggest that as much as 21% of GTS heritability can be explained by genotypes with a MAF (minor allele frequency) between 0.001 and 0.05 (Davis et al. 2013). Recently, a whole-exome sequencing study indicated a role of the rare variant burden in 13 families with TD history (Cao et al. 2021). Still, the role of rare, and particularly non-coding variants, particularly with MAF < 0.001 remains largely unexplored for GTS and other TDs.

To verify the above conjecture, we investigated whether an oligogenic additive model could be used to distinguish healthy individuals from GTS and other TDs subjects. Whole-genome sequencing (WGS) was used to analyze known and identify novel variants in 84 genes previously indicated with varying levels of evidence, to be associated with GTS and other TDs. The search window also encompassed 20,000 bp of both flanks of the gene to identify variants located both in the gene itself and in distant regulatory regions as well. In the first phase of the analysis, ten genes were selected for the oligogenic model which was then verified on GTS patients from 23 families, their healthy relatives and a group of whole genome sequences of 100 unrelated Polish sportsmen free of any neurological disorders. This oligogenic score was then also calculated for individuals with non-GTS TDs and showed that, on average, their score resides below GTS but above healthy individuals.

This is, to our knowledge, the first statistical model using WGS data, including mostly non-coding and many rare variants, to predict GTS/TD risk. The obtained results were additionally compared with a polygenic risk score (PRS) based on the Psychiatric Genomic Consortium (PGC) GTS GWAS study (Yu et al. 2019). Overall, our results provide novel insights into the genetic architecture of GTS and other TDs and contribute evidence that they reside on a common genetic spectrum.

## Methods

### Patient selection and diagnostic criteria

All patients were recruited from a single Outpatient Clinic and were personally reviewed and evaluated by the same clinician well-experienced in tic disorders (PJ). The patients had been referred to the Clinic by a general neurologist and psychiatrist due to problematic diagnoses or tics refractory to treatment or sought medical advice on their own because of troublesome tics. The study was designed as a one-time registration study, as patients were registered in the database only once, and no additional clinical data obtained in follow-up visits were included in the analysis. In case of a positive family history, DNA was collected from affected relatives and healthy members of the proband’s family.

The patients were evaluated for GTS and other TDs (Chronic Motor Tic Disorder, Chronic Vocal Tic Disorder, Provisional Tic Disorder, Other Specified Tic Disorder and Unspecified Tic Disorder) according to the Diagnostic and Statistical Manual of Mental Disorders criteria valid at the time of evaluation (DSM-IV-TR, DSM-5). All patients were systematically interviewed with the aid of a semi-structured interview consisting of demographic and clinical data. This schedule was based on the TIC (Tourette syndrome International database Consortium) Data Entry Form developed by Freeman et al. (Freeman et al. 2000), in which the investigator (PJ) participated and subsequently used this form in clinical practice.

The prevalence of the most common comorbid disorders encountered in GTS was evaluated based on the above semi-structured clinical interview. The disorders listed in this interview included: ADHD, OCD, depression, anxiety disorder (different forms of phobias, panic disorder, generalized anxiety disorder, and separation anxiety disorder), Oppositional Defiant Disorder, Conduct Disorder, and Autism Spectrum Disorder. The list of obsessions and compulsions included in the Yale-Brown Obsessive Compulsive Scale (Y-BOCS) was used to establish the clinical spectrum of OCD. All patients were questioned thoroughly about all the symptoms included in the DSM as the diagnostic criteria of comorbid disorders mentioned above. Diagnoses of mental disorders from psychiatric clinics established before our evaluation were accepted and included in the analysis. The diagnosis of Autism Spectrum Disorder was made in specialized clinics. Children and adolescents with more complex psychopathology were assessed with the M.I.N.I. International Neuropsychiatric Interview for Children and Adolescents. Patients with severe psychiatric comorbidities were referred to a psychiatrist to confirm the diagnosis.

Tic severity was measured using the Yale Global Tic Severity Scale (YGTSS) on the day of a patient’s evaluation (Leckman et al. 1989). An additional diagnosis of severe GTS in adult patients was also assessed. In contrast to children and adolescents, for whom most of the information was provided by their parents, adults reported the symptoms themselves. Tics were qualified as severe if at any time they disrupted normal daily activities (e.g. led to repeating grades at school, job loss, or physical injuries), caused a significant deterioration of life quality, or required prolonged pharmacological therapy. Tics were also qualified as severe if the Yale Global Tic Severity Scale-Total Tic Score (YGTSS-TTS) was ≥ 35 points (range: 0-50) at the time of clinical evaluation (Leckman et al. 1998).

Overall, each patient was assigned to one of the following groups: GTS, non-GTS TDs, or healthy controls. In the GTS group, an additional subgroup of severe tics was distinguished according to the criteria described above.

### Whole-genome sequencing

Genomic DNA of 186 patients (eight samples were later excluded at various stages during extensive data quality control) was extracted from peripheral blood leukocytes using a standard salting-out procedure (Miller, Dykes, and Polesky 1988) or from saliva collected with the Oragene DNA Self Collection Kit and using the Prep IT L2P Purification Kit (DNA Genotek Inc., Ottawa, Ontario, Canada). Whole-genome sequencing (WGS) was performed by Novogene (Beijing, China) according to the following protocol: sequencing libraries were generated using the NEBNext Ultra II DNA Library Prep Kit for Illumina (New England Biolabs, the UK) following the manufacturer’s recommendations. Genomic DNA was randomly fragmented to 350 bp on average with a Bioruptor and DNA fragments were size-selected with sample purification beads. The selected fragments were then end-polished, A-tailed, and ligated with a full-length adapter. The fragments were then filtered with the beads again. Finally, the libraries were analyzed for size distribution on an Agilent 2100 Bioanalyzer, quantified using real-time PCR, and paired-end sequenced on an Illumina high-throughput HiSeq X Ten sequencer.

### WGS data preprocessing

Fastq files were processed with the Intelliseq Germline Pipeline (https://gitlab.com/intelliseq/workflows, https://flow.intelliseq.com/) built with Cromwell (https://cromwell.readthedocs.io/en/stable/). The fastq files were assessed for quality with FastQC. The files were then aligned to the Broad Institute Hg38 Human Reference Genome with GATK 4.0.3. Duplicate reads were removed with Picard and base quality Phred scores were recalibrated using the GATK covariance recalibration. Variants were called with the GATK HaplotypeCaller to give genomic variant calling files (gvcf)

### Additional control samples and synthetic controls

From this step all the analyses were run within the Hail library (v 0.2.113). All the code used in this project is available in the project’s github repository (https://github.com/ippas/imdik-zekanowski-gts/tree/master/preprocessing/oligogenic-mod el). To ensure the robustness of the results two additional control groups were used. In the discovery phase we included synthetic controls based on a database of allele frequencies of the local (Polish) population (Kaja et al. 2021). Synthetic samples were genotyped with MAF of each of the variants in the database as probabilities of assignment of a given allele. The sex distribution of the simulated samples reflected the sex distribution in the study group. The Y chromosome was excluded from the analysis and simulated males were assigned homozygous genotypes in non-PAR X regions. Each locus was drawn independently and the final synthetic dataset consisted of 25 groups of 100 samples. The control group was a cohort of 100 Polish elite sportsmen free of any neurological disorders (data was available locally as .gvcf files and processed in the same way as files for this study).

### Merging of the genomic .vcf files, variant filtering and annotation

Gvcf files (study samples and local controls) were imported to hail’s native vds format. At this step the dataset was filtered to contain only loci within +/-20kb windows from protein-coding genes and QC was performed. First, variants were filtered to exclude repeated and low-quality sequences (UCSC RepeatMasker track). Multiallelic variants were split and only loci with more than 90% gnomAD (v3) samples with a DP > 1 were kept for further analysis (Karczewski et al. 2020). Variant quality statistics were then computed separately for the GTS and Polish sportsmen cohorts. Only sites with mean DP > 5, mean GQ > 40, Hardy-Weinberg equilibrium test p-value > 1e-15, no more than 8 samples with DP < 3 and no more than 40 samples with GQ < 30 were kept. Six samples were removed during this filtering process. Variants were annotated with gnomAD (v3; https://gnomad.broadinstitute.org/) (Karczewski et al. 2020), combined annotation dependent depletion (CADD) scores (Rentzsch et al. 2019), and human phenotype ontology (HPO) phenotypes (Köhler et al. 2021). For a general overview of the factors determining genetic variance, HWE-normalized Principal Component Analysis (PCA) was run on randomly sampled 20% (∼600k) of all high quality variants after the filtering. Two samples that were outliers (more than 10 SDs from the mean on any of the first three Principal Components - PCs) were excluded from further analyses.

### Burden tests for gene selection

In the first part of the analyses GTS patients outside families (n = 37) were compared with synthetic controls based on a database of WGS allele frequencies in the Polish population (Kaja et al. 2021). To ensure robustness of the results 25 comparisons were run at this step (GTS patients vs each one of 25 groups sampled from 2500 synthetic controls). First, for each of the 25 comparisons, PCA was conducted on 20% of randomly sampled genotypes (∼280 k variants for each subset). Then, preliminary sequence kernel association test (SKAT) tests in logistic mode (Wu et al. 2011) for all protein coding genes with 20000 kb flanks were run to find the optimal number of PCs for correction. CADD scores were used as weights for the SKAT test. Four PCs were chosen, as whole-genome results corrected for 4 PCs had the mean p-value inflation factor closest to 1. Still, the median lambda GC was 0.73 - a deflation of p-values likely due to low power. Then SKAT tests were run for variants in and in the vicinity of 84 genes previously implicated in GTS again as 25 comparisons (unrelated GTS patients vs each of the synthetic control groups). The resulting p-values were then averaged from 25 comparisons and only genes with an average p-value < 0.05 and no p-value > 0.25 were chosen for the oligogenic classifier.

### Oligogenic classifier construction and optimisation

The top 10 top genes from the SKAT test results were used for oligogenic risk models of GTS. For each gene and sample, the number or weights of variants that passed chosen thresholds was summed and standardized with the mean and SD of the whole study sample. The per-gene scores were then summed for the final oligogenic score for each sample. A total of 12 classifiers were tested. These differed in CADD score threshold (0,5,10), MAF filter applied: either MAF < 0.0001 in the GnomAD non-Finnish European (NFE) population or no MAF filter and in the application of weights to variants (CADD as weights or no weights). The models were run on a group comprising GTS and healthy individuals from 23 families and 100 Polish elite sportsmen. The AUC-ROC of the classifiers was calculated based on the sliding of the threshold of the oligogenic score. For the top classifiers, to estimate type I error, we used a Monte Carlo feature selection approach. In this approach features (genes) to build models were selected randomly in each iteration and AUC was calculated. The p-value of the AUC for the original model was evaluated using the distribution of AUC values of random models. Random gene sets were selected from a list of human protein-coding genes that are assigned to at least one Gene Ontology term to be comparable with the selected gene set. The procedure for the random genes was identical to the one for the evaluated models.

### Application of the oligogenic model to all groups

For the best-fit model defined as the highest AUC model, we applied the oligogenic scores to all groups used in the previous analyses stratified according to tic severity, including a group of individuals from the family cohort that were diagnosed with non-GTS TDs not used in the previous analyses. All groups were compared with pairwise t-tests without correction for multiple comparisons. The same procedure was applied to different groupings (by sex and comorbidities). Percentages of samples assigned as GTS or not by the oligogenic model were calculated based on the optimal point of the AUC curve of the best-fit model defined as the point with the highest sensitivity and specificity sum. All variants included in the risk model were characterized regarding their position in relation to the gene, population allele frequency predicted molecular consequences, and predicted pathogenicity based on their annotations.

### PGC GWAS signal overlap and PRS analysis

GTS GWAS data were obtained from the Psychiatric Genomics Consortium (Yu et al. 2019) (https://www.med.unc.edu/pgc/download-results/). The SNP coordinates were translated from Hg37 to Hg38 coordinates with the Hail liftover function. SNPs within the classifier genes and in 20,000 bp flanks of the genes were investigated in terms of their p-values from GWAS using quantile-quantile (qq) plots. Random gene sets of 10 genes and their lambda GC inflation factors on qq plots were used to determine whether these p-values showed a distribution different from that for the whole genome. The PRS was developed based on GWAS results using polygenic package version 2.3.13 (https://github.com/intelliseq/polygenic). Briefly, the procedure includes filtering out indels, filtering out ambiguous variants with complementary alternative alleles (A/T or C/G), converting to genomic ref/alt allele names, liftover to Grch38, filtering for p-value threshold (10e-50, clumping, converting odds ratio to beta coefficients and assigning gene symbols. The PRS model was then stored in yml format. To calculate PRS genotyping of gvcf files was executed using the GATK GenotypeGVCFs v4.1.7.0 tool. Regions annotated as having alternate loci in GRCh38 were excluded. PRS scores were calculated using the polygenic 2.2.4 tool. PRS-scores were compared with pairwise t-tests.

## Results

### Description of the study group and overview of the analyses

The study group comprised GTS patients and their families from the Polish population as well a group of 37 unrelated GTS patients. A group of 100 Polish athletes free from any neurological and psychiatric disorders was included as control. All the included family members of GTS patients were diagnosed towards GTS and other TDs. Each subject was assigned to one of the following sub-groups: healthy, severe GTS, GTS, and other tic disorders. Out of 278 subjects included in the final study group, 37 were unrelated individuals, including 35 adults diagnosed with GTS with severe tics. The remaining subjects (n=141) came from 23 families with 3 to 14 members each (median 6 members), and a cohort of Polish elite sportsmen (n=100). Overall, 55 out of the patients included in the study were diagnosed with comorbidities, including 11 with ASD, 24 with OCD and 19 with ADHD. Table 1 presents the distribution of basic and additional diagnoses.

**Table 1.** General characteristics of study groups. GTS - Gilles de la Tourette syndrome, non-GTS TD - tic disorder other than GTS, ASD - Autism Spectrum Disorder, OCD - Obsessive-Compulsive Disorder, ADHD - Attention-Deficit/Hyperactivity Disorder Although multiple genes have been proposed to be associated with GTS and TDs most of them are only backed up by limited evidence (Levy, Paschou, and Tümer 2021; Lin, Tsai, and Chou 2022) and the role of rare and non-coding variants in these genes is uncertain. To investigate the possible oligogenic model of GTS we constructed a list of genes (n=84) from diverse genetic studies on GTS (association studies, linkage analysis, GWAS, WES/WGS) offering a varied quality of evidence (Table 2). We then investigated variants in and near these genes in WGS data. Overall, the study consisted of four steps: first, top genes for the oligogenic score were selected based on a burden test, then the risk model was constructed and applied. Finally, we overlapped the score with the common variant GWAS-based PRS (Figure 1).

| diagnosis | number of patients | sex | additional details | comorbidities |
| --- | --- | --- | --- | --- |
| **discovery group (unrelated patients)** |  |  |  |  |
| GTS | **37** | 6 female,<br>31 male | including 35 adults<br>with severe tics | 4 ASD, 14 OCD, 17<br>Depression, 16<br>ADHD, 20 Anxiety<br>Disorder |
| **family group** |  |  |  |  |
| GTS | **46** | 14 female,<br>32 male | including 3 adults<br>with severe tics | 5 ASD, 6 OCD, 6<br>Depression, 3<br>ADHD, 7 Anxiety<br>Disorder |
| non-GTS<br>TD | **40** | 14 female<br>26 male |  | 1 ASD, 4 OCD |
| healthy<br>controls<br>without tics | **155** | 37 female<br>118 male | including 55<br>subjects from GTS<br>families | 1 ASD, 1 Depression |

**Table 2.**
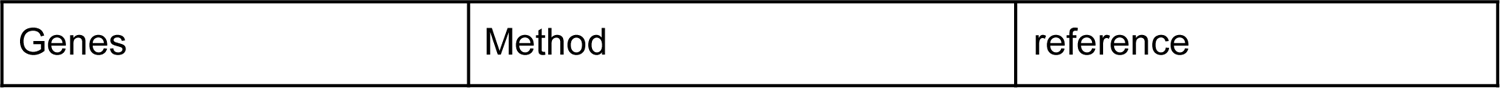
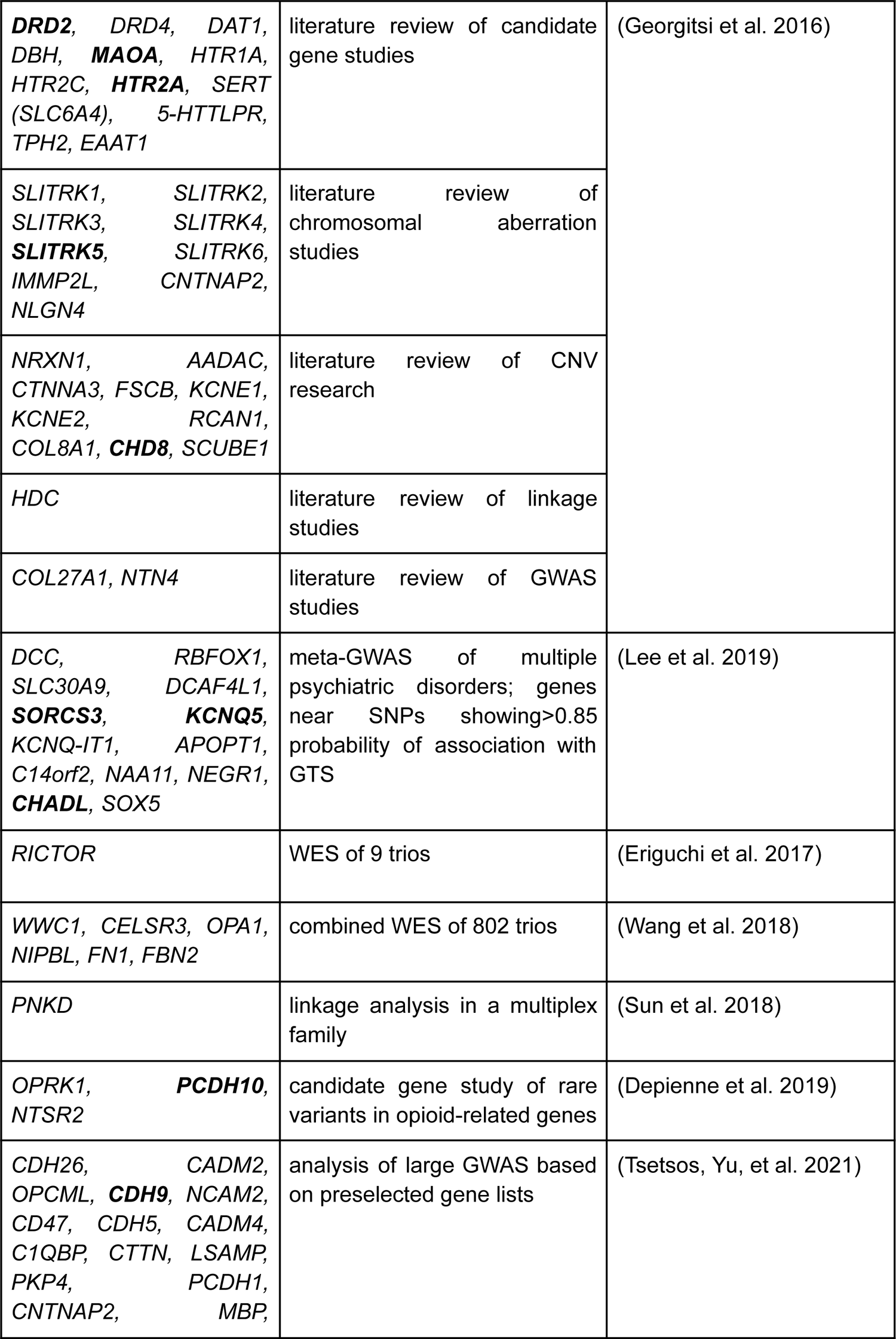
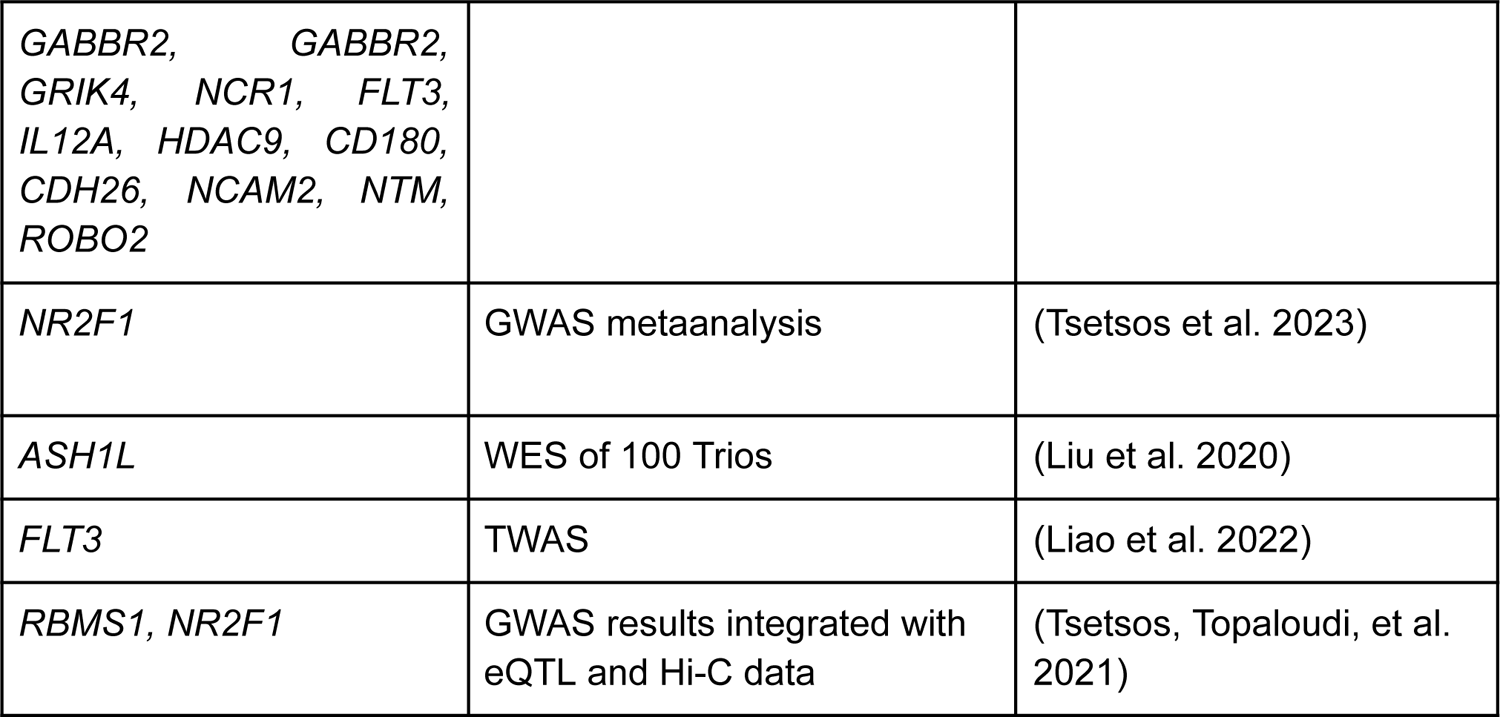
List of genes potentially related to GTS used for oligogenic models of GTS. Gene symbols in bold font indicate genes that were included in the final developed model.

### WGS data description

The core dataset of the study consists of variants within and in the vicinity (+/- 20000 bp) of protein-coding genes. After quality control filtering, in the 278 samples there were ∼6.5 million single nucleotide variants (SNVs, exact n = 6,514,756). 5.2 million variants were observed in the GTS cohort (n = 178), while 4.4 had non-reference calls in the cohort of Polish sportsmen (n = 100). Of all genetic variants, 4.3 million were in introns, about 765,000 variants were in exons, 427,000 in UTRs, and the rest were located in intergenic regions. Per-group allele frequencies in each genotyped locus in the core dataset including 20,000-bp 5’ and 3’ flanks are reported in Supplementary Material (Supplementary Table S1). The per sample call rate in the final dataset was above 99.5% for all samples. The mean phred-scaled genotype quality was 75.9 and the mean coverage 29.7 (Supplementary Figure S1). PCA of the whole dataset (with the synthetic samples included) revealed that belonging to the largest families was the factor that determined the major PCs (Supplementary Figure S2).

### Feature selection - identification of genes in which variants contribute to GTS risk

In order to select genes that will be used for the oligogenic model we applied the SKAT test, which accounts for the contribution of both common and rare variants and SNP-SNP interactions (Wu et al. 2011). To focus on the damaging variants Combined annotation-dependent depletion (CADD) scores predicting the pathogenicity of both coding and non-coding variants were used as weights for the SKAT. The higher the predicted pathogenicity of the variant the larger its weight. CADD leverages a range of variant annotations (including SIFT and PolyPhen) and, importantly, provides scores for non-coding variants as well (Rentzsch et al. 2019). In this step, the discovery samples of unrelated GTS patients (n = 37), mostly with severe tics that persisted into adulthood (n = 35) were compared with synthetic controls. To ensure robustness of the results GTS patients were compared with synthetic controls 25 times, each time with a different group of 100 synthetic controls. Out of the 84 comparisons, 10 genes were selected (Table 3, full results available in Supplementary Table S2). We observed stability of the result with various control groups as 5 genes *CHADL, DRD2, MAOA, PCDH10* and *HTR2A* had p-values < 0.05 in all or all but one comparisons, and the remaining 5 genes (SLITRK5, SORCS3, KCNQ5, CDH9 and CHD8) with p-values < 0.05 in at least 75% of comparisons.

**Table 3.** Selection of genes with possible overrepresentation of rare, damaging variants in unrelated GTS patients. Results of the SKAT test (unrelated GTS vs synthetic controls based on population-specific allele frequencies). To ensure stability of the results a group of 2500 synthetic controls was divided into 25 groups of 100 synthetic samples. Here we display averaged statistics, full statistics are available in supplementary Table S2. Genes with average nominal p-values < 0.05 are shown. These 10 genes were selected for the oligogenic model.

| gene symbol | number of variants | SKAT p-value |
| --- | --- | --- |
| CHADL | 96 | 0.00119 |
| MAOA | 5 | 0.00337 |
| DRD2 | 215 | 0.00422 |
| PCDH10 | 234 | 0.00709 |
| SORCS3 | 1327 | 0.0201 |
| KCNQ5 | 1099 | 0.0262 |
| CDH9 | 447 | 0.0263 |
| SLITRK5 | 135 | 0.323 |
| CHD8 | 146 | 0.0364 |
| HTR2A | 245 | 0.0380 |

### Oligogenic risk model construction and evaluation

The first step allowed us to select top GTS candidate genes with possible overrepresentation of damaging variants in GTS patients. Next, a limited number of classifiers based on the 10 top genes selected in the discovery phase was created. We applied three different CADD thresholds as cut-off values for variant inclusion (0, 5, or 10), two options for MAF cut-off (0.0001 in non-finnish European population or no cut-off), weighted (by CADD) and unweighted models. Oligogenic scores were standardized per gene and summed in each case. This created 12 oligogenic models, which were applied to a task of distinguishing GTS patients from their healthy relatives and a group of 100 unrelated controls from the same population. The best-fit model was based on all variants with CADD scores detected in 10 genes normalized per-gene with the Area Under the Curve of the Receiver Operator Characteristics (AUC-ROC) of 0.62 and the second best was a model that only included variants with MAF < 0.0001 AUC-ROC 0.60 (Figure 2A). To control for spurious findings, for each of the top classifiers we constructed 1000 random control classifiers with the same conditions, but with randomly selected features (genes) in a Monte Carlo approach (Figure 2B, Supplementary Figure S3).

**Figure 1.**
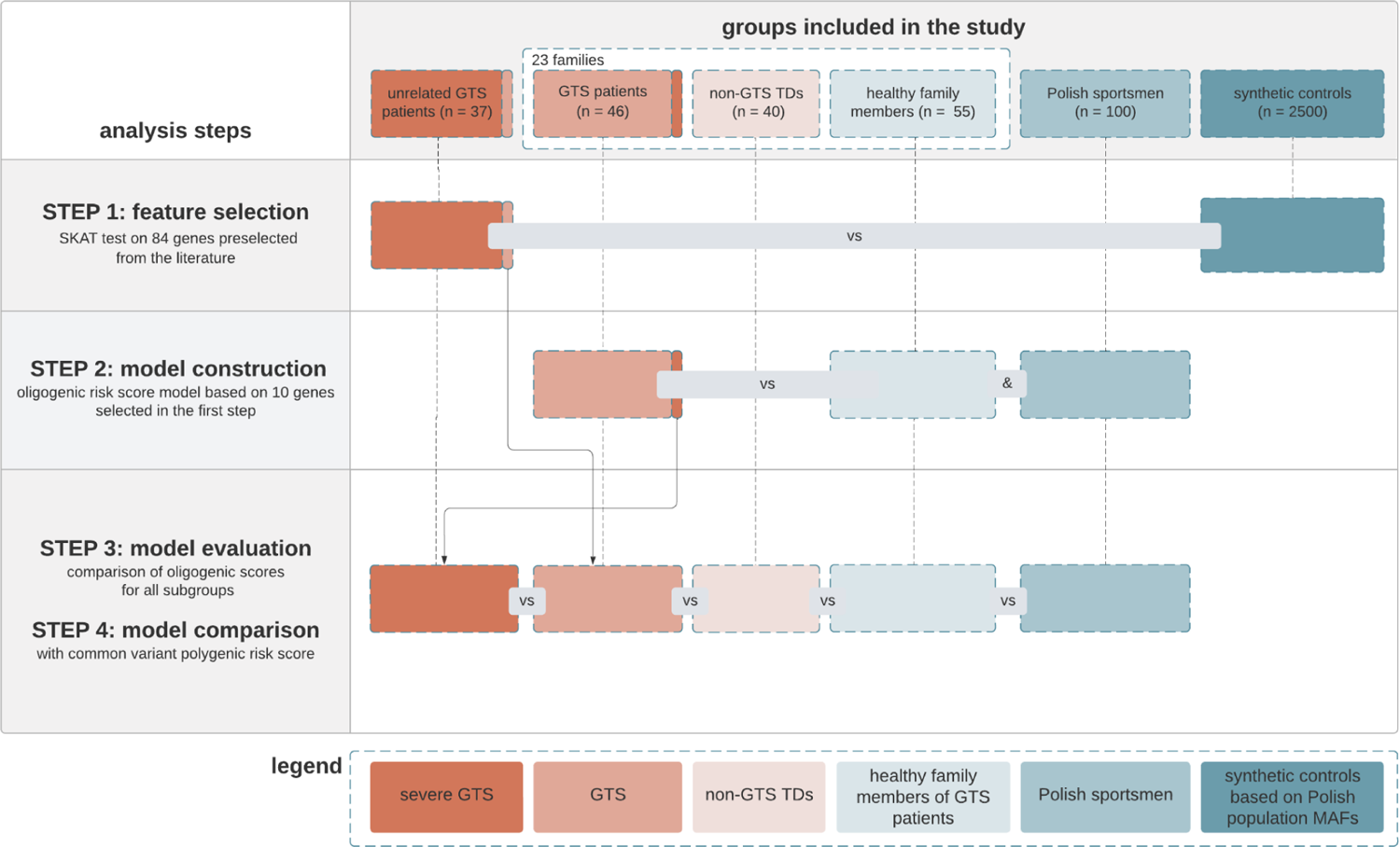
Overview of the study and groups analyzed at each step. The study included 5 groups of samples and 1000 synthetic controls. In the feature selection set unrelated GTS patients were compared with synthetic controls (STEP 1). In the model construction part, GTS individuals were compared with healthy family members and a group of athletes from the same population (STEP 2). Then comparisons of oligogenic scores between five groups including a group of patients diagnosed with non-GTS TDs were performed (STEP 3). The oligogenic model, predominantly based on rare variants, was then compared with common variant PRS (STEP 4).

**Figure 2.**
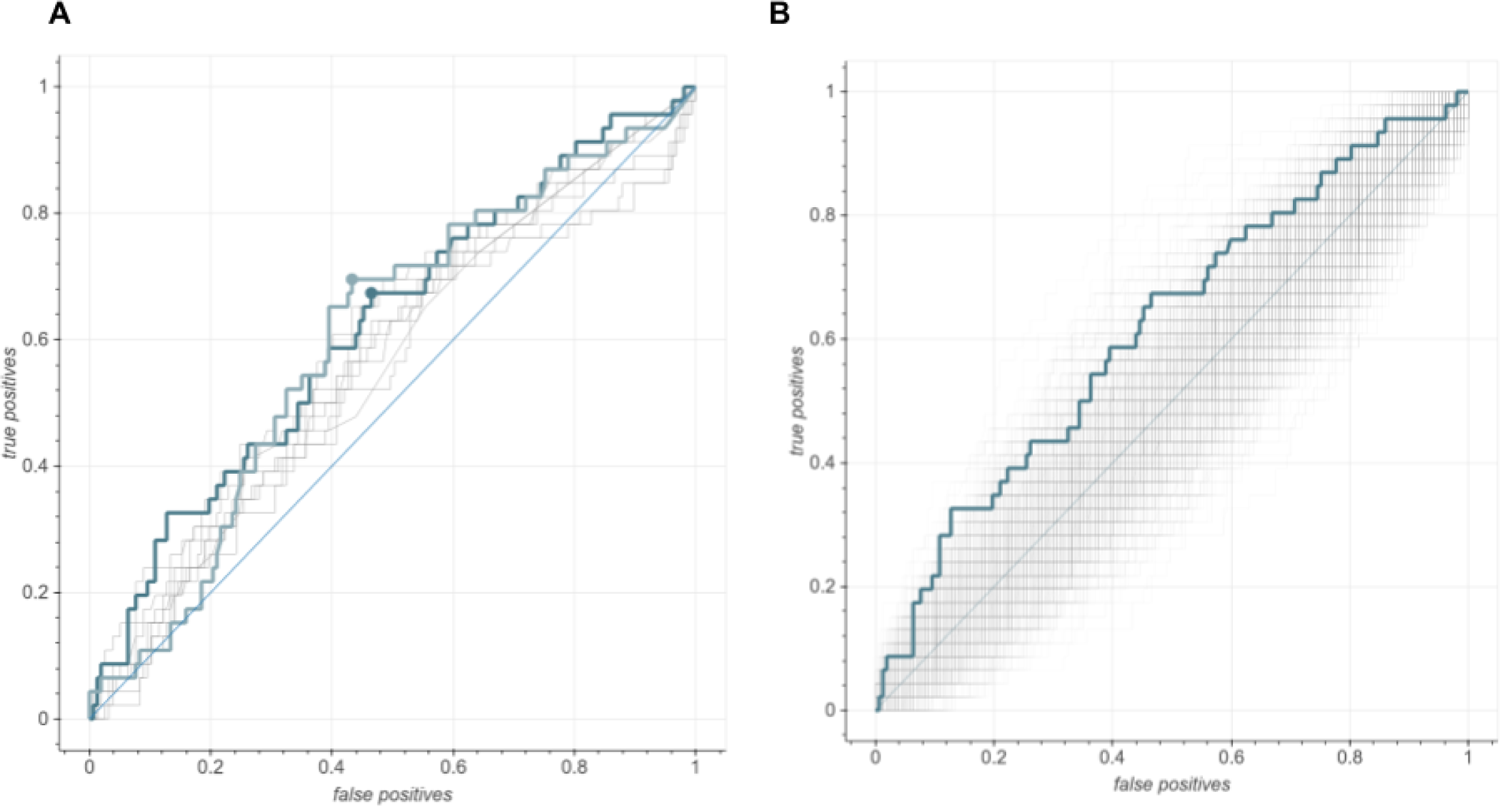
Comparison of the performance of tested classifiers of GTS risk. Graphs show ROCs of the best-fit GTS risk model and other tested classifiers. **A.** Comparison of models based on the top 10 genes from the previous steps with different CADD thresholds, MAF filter and weights. The dark blue thick line represents the best-fit classifier taken as a GTS risk model (AUC = 0.62 95% CI = [0.52 - 0.71]), the light blue thick line is for the second best model - model with the highest sum of sensitivity and specificity (AUC = 0.60, 95% CI = [0.51 - 0.70]). Circles show the point with the most optimal assignments (The best fit model: false positive percentage: 46.5%, true positive percentage: 67.4%; the model with the highest optimal point: false positive percentage: 43.3%, true positive percentage: 69.6%) the rest of the tested classifiers are shown with thin gray lines **B.** Best-fit model from A compared with 1000 control classifiers (thin gray lines) based on sets of random genes and the same parameters. x-axes: fraction of false-positive results; y-axes: fraction of true positive results.

### Evaluation of the oligogenic risk model

After the best-fit model was selected we applied the oligogenic score to all of the groups of the study. Crucially, this part included a group of individuals with non-GTS TDs. These samples were not used neither in the feature selection nor in the model construction steps. The analysis revealed that healthy samples had the lowest mean oligogenic score, followed by patients with non-GTS TDs, then GTS and GTS with severe tics (Figure 3). As the groups of the study were unbalanced with regards to sex we evaluated the oligogenic score per sex and found no difference (Supplementary Figure S4). Furthermore we investigated if the score is associated with any of the comorbidities in the GTS cohort (Supplementary Figure S4). The only statistically significant difference wes between the groups with and without anxiety disorder. This may be explained by the fact that the majority of patients with this comorbidity also had severe GTS.

**Figure 3.**
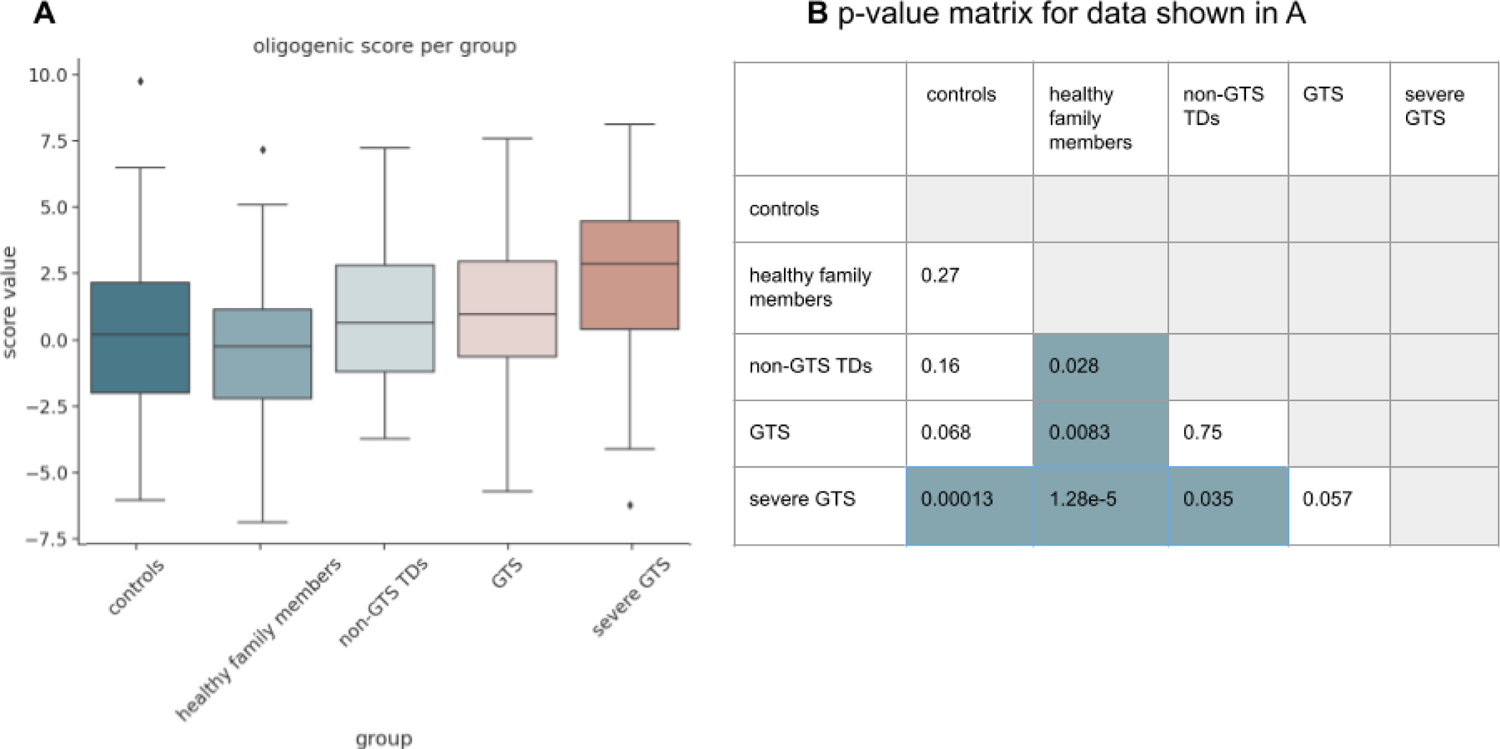
Comparison of the oligogenic risk score between all groups included in the study. **A.** Boxplot of oligogenic scores for each of the groups included in the study. Horizontal lines show the median value, error bars are interquartile ranges. **B.** P-value matrix of pairwise t-tests performed for data in (A). Cells with p-values < 0.05 are marked in blue. Detailed t-test results and the Shapiro-Wilk tests of normality results are supplied in Supplementary Table S3.

To further characterize the genetic variants present in the oligogenic model we analyzed them in terms of their putative functional consequences. Out of the 7654 variants included in the best-fit model, 5341 had non-reference calls in GTS patients, 3868 in non-GTS TD patients and 5997 in healthy individuals (which was the largest group in the study). Exactly 3807 variants were overrepresented in GTS patients. Over 1300 of the variants were present in the *SORCS3* gene, while *MAOA* had only 5 SNPs. Variants were checked for MAF in the NFE population in gnomAD and the median MAF of all of the variants was 0.02 (Figure 4A). 760 variants had allele frequencies below 0.0001 and this rare-variant subset was the basis for the second-best model (AUC = 0.60). The variants overrepresented in the GTS cohort showed a range of functional consequences, being mainly intronic and other non-coding variants. About 1% of variants were missense, and no protein truncating variants were included (Figure 4B). Finally, we investigated how the chosen oligogenic model performed per-family where the optimal point of the ROC-AUC curve was determined as the cutoff. Figure 4C shows such an assignment for two largest families of the group: a family with 11 members (family F - family 1 in Figure 4C) and the largest family in the dataset (family W - family 2 in Figure 4C), with 14 members. For family F the true positive rate was 100% (8/8 - for classification individuals with non-GTS TDs are considered cases) and the false positive rate was 33% (1/3). For family W the false positive rate was 33% (2/6) and the true positive rate 62.5% (5/8). In both families individuals with GTS and other TDs have, on average, more variants from the list included in the best classifier Supplementary Table S4 provides assignments by the model for all families.

**Figure 4.**
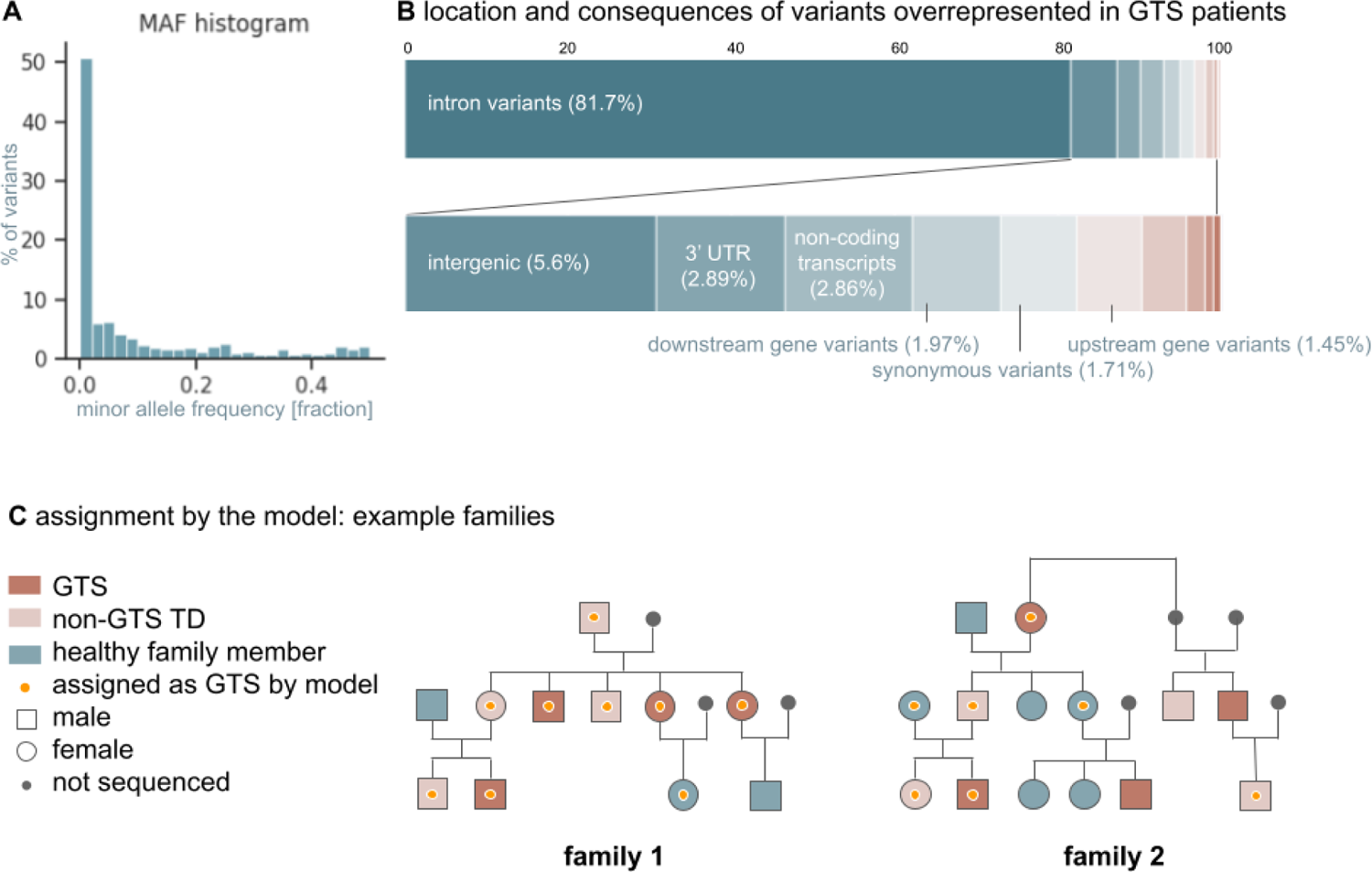
Characteristics of genetic variants included in the GTS risk model and example assignments. **A.** Distribution of minor allele frequencies (MAF) of 7654 variants in non-Finnish Europeans in the gnomAD database; **B.** Functional consequences/localisation of variants overrepresented in GTS samples according to Ensembl Variant Effect Predictor (VEP) most severe consequence assignment for each variant (McLaren et al. 2016). Only variant types that made up over 1% are described in the figure. Other variants and their percentages are: missense (0.99%), variants in regulatory regions (0.42%), 5’ UTR variants (0.18%), splice region variants (0.16%), splice donors (0.03%); **C.** Classification of members of two largest families of the studied group by the GTS risk model. Orange dots indicate individuals assigned to the GTS and non-GTS TDs group by the risk model at the optimal cut-off threshold.

### Comparison of the oligogenic model with a common-variant polygenic risk score

To date, the largest publicly available GTS GWAS dataset is the study carried out by the PGC (Yu et al. 2019). It has been shown that the PRS based on this GWAS predicted tic presence in another cohort and explained about 0.5% of the variance in tic presence in the external validation dataset (Abdulkadir et al. 2019). Out of the 10 genes included in the oligogenic model, three genes *SORCS3, KCNQ5 and CHADL* were selected based on a meta-analysis of multiple GWAS for psychiatric disorders (Lee et al. 2019). This meta-analysis did include the GTS PGC dataset. We, therefore, first interrogated whether the common variant background investigated by GWAS overlaps with genes included in the model. After filtering the GWAS summary statistics for SNPs within the 20 000 bp windows and each gene loci 5929 were found, including 10 SNPs with p-values < 10-e5. All the SNPs with p-values < 10e-5 were in the CHADL gene locus. Overall, the lambda GC p-value inflation factor of all 5929 SNPs found within and in the vicinity of the 10 genes in our model was 1.33, which does indicate some inflation from the overall dataset (1.08) (Figure 5). However, after performing a false-positive rate control by checking the lambda GC for 1000 random sets of 10 genes we found that 1.33 was in the 79th percentile of all of the sets (empirical p-value = 0.21). Therefore we concluded that there is no significant overlap between the WGS-based signal included in our model and the common variant GWAS signal, apart from the *CHADL* locus.

**Figure 5.**
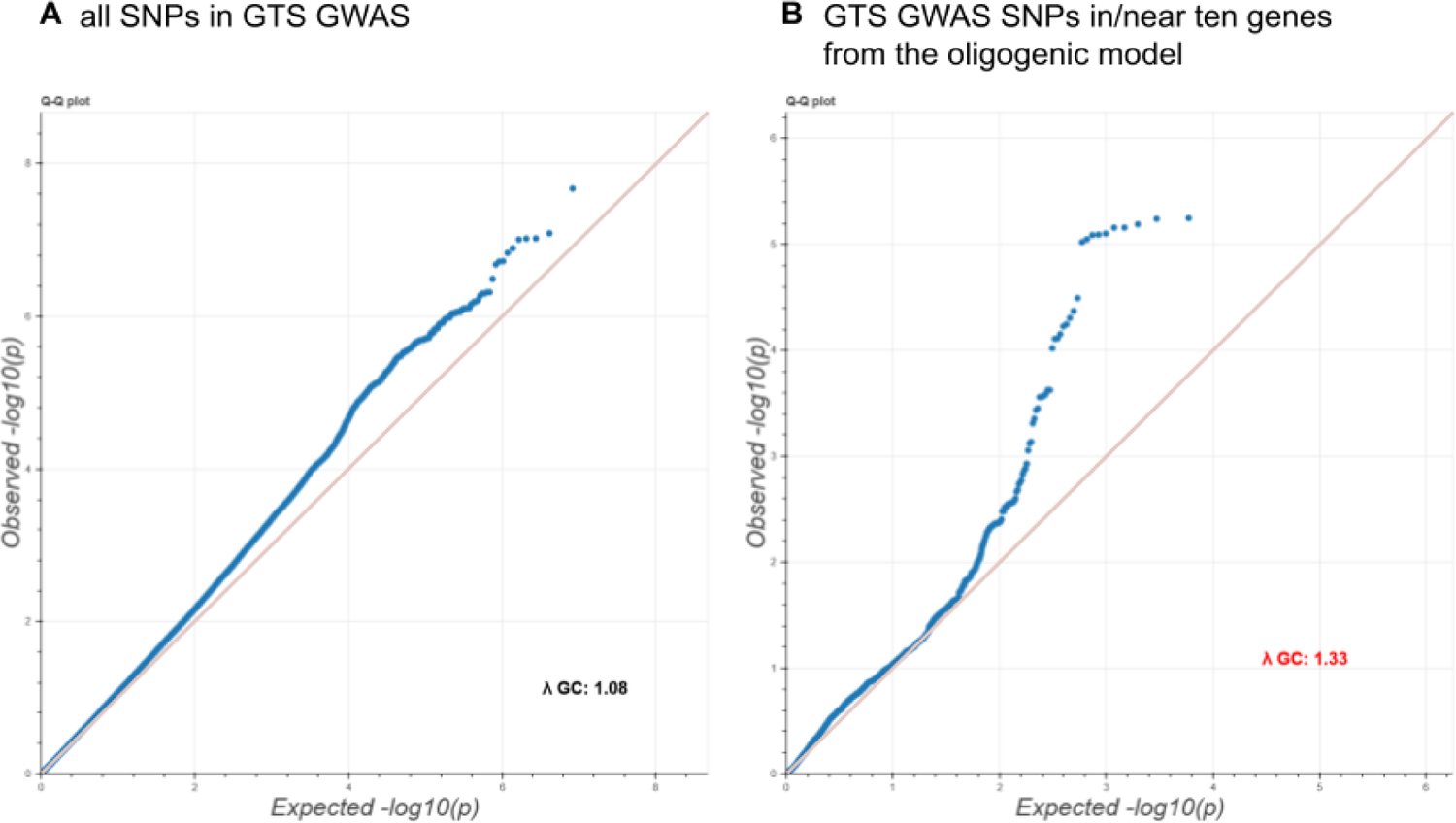
Overlap of common-variant GTS associations with the oligogenic model. Figure shows quantile-quantile (qq) plots of expected (x-axis) versus observed (y-axis) p-values. **A.** Qq plot for all of the SNPs included in the PGC GWAS dataset (lambda GC inflation factor 1.08). **B.** Qq plot for 5929 SNPs investigated by the PGC GWAS and found within the +/- 20kb flanks of the loci of ten genes included in the oligogenic model (blue dots, lambda GC inflation factor 1.33)

As the oligogenic model and common variant signal appeared to arise only from partially overlapping loci, we applied GWAS-based PRS to all of the samples to compare the values with the oligogenic model. GWAS SNPs below the 10e-5 p-value threshold were included in the model as this was the lowest p-value threshold with over 10 variants. This analysis revealed that for the same task as the initial oligogenic model the common-variant PRS model performed at chance level (AUC-ROC = 0.47). We additionally assessed classification performance of the ten-gene oligogenic model when it was augmented with the common variant PRS and found that it worsened the model performance (AUC-ROC = 0.53, Figure 6A). PRS scores for each GTS subgroup were also compared (Figure 6B). The control group of Polish sportsmen had the lowest median value (−0.69), and the severe GTS subgroup the highest (−0.41). The groups from the family cohort had in-between median values and were not significantly different from each other. In pairwise tests, the only nominally statistically significant result was between the group with non-GTS TDs and the sportsmen control cohort (p = 0.035).

**Figure 6.**
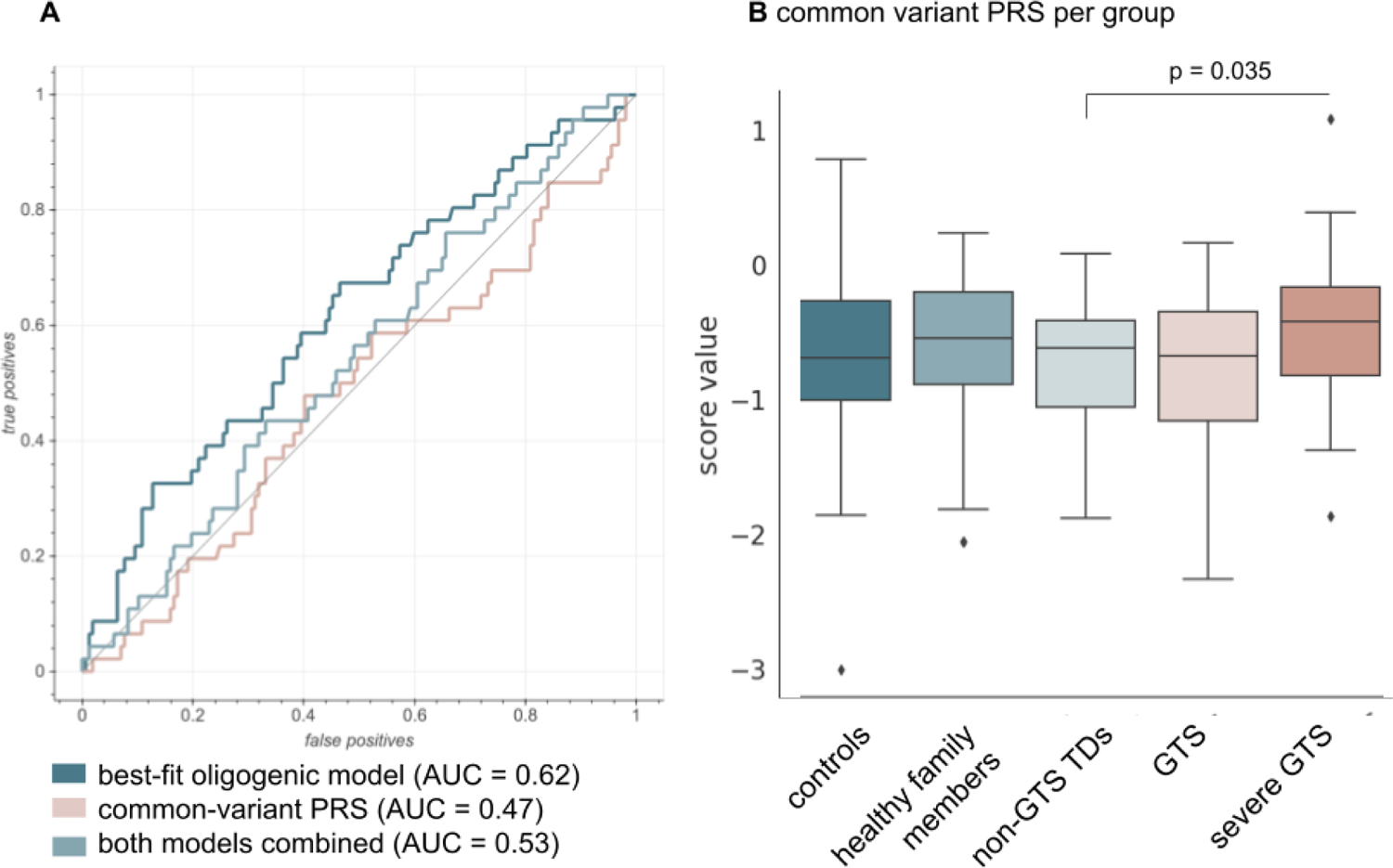
Comparison of the oligogenic model with GWAS-based PRS. **A.** ROC-AUC graphs for the best-fit oligogenic model, the PRS, and the combined model for the task of distinguishing healthy individuals from GTS patients. **B.** Boxplots for per group PRS. Groups were compared with pairwise t-tests. Nominal p-values <0.05 are reported in the graph.

## Discussion

Studies published to date indicate that GTS has a polygenic background and that rare variants may have a high impact on GTS etiopathology (Lee et al. 2019; Wang et al. 2018; Willsey et al. 2017). It has also been suggested that the same genetic factors contribute to various developmental TDs, including GTS, with a higher burden causing more severe disease (Robertson et al. 2017). Here we proposed an oligogenic risk model of GTS which supports this increasing variant burden hypothesis. The main conclusion of our study is that non-GTS tic disorders, childhood typical GTS (with more severe tics in childhood and subsiding in adolescence and adulthood), and severe GTS that persists into adulthood share genetic background with different burden values. Patients with severe GTS have the highest genetic risk burden indicated by both common and rare variants. We additionally report novel rare and non-coding variants in and near GTS-risk associated with the disease.

Our study was performed on a group of 278 Polish subjects with whole-genome sequencing. This group included GTS patients, their healthy relatives, and an additional control group that consisted of elite sportsmen from the same population. Uniquely, our study also involved patients with severe GTS, meaning patients with troublesome tics that persisted into adulthood. To further augment the analyses we used simulated samples based on a database of allele frequencies of the general Polish population (Kaja et al. 2021). The presented oligogenic risk model was developed in two steps and is based on variants in and in the vicinity of ten genes; *CHADL*, *DRD2*, *MAOA*, *PCDH10*, *HTR2A*, *SLITRK5*, *SORCS3*, *KCNQ5*, *CDH9,* and *CHD8*. Although the genes included in the risk model have previously been claimed to be associated with GTS, for most of them no such link was confirmed by large-cohort studies. These genes can be divided into two groups: those based on candidate gene studies (*DRD2*, *MAOA*, *PCDH10*, *HTR2A*) and those from more high-throughput analyses such as GWAS or CNV research (*CHADL*, *SLITRK5*, *SORCS3*, *KCNQ5*, *CDH9,* and *CHD8*). These genes were previously supported by various evidence levels. As an example, two rare (MAF < 1%) variants in *PCDH10* that possibly lead to a loss of protein function have been detected in GTS patients, while *SORCS3, KCNQ5* and *CHADL* were implicated in a large meta-analysis of multiple GWAS (Depienne et al. 2019; Lee et al. 2019). Their biological roles are diverse and include genes which products are involved in: monoamine neurotransmission (*DRD2*, *MAOA*), the opioid system (*PCDH10*), serotonin neurotransmission (*HTR2A*), regulation of neurite and synapse development (*SLITRK5, CDH9, CHD8*), synaptic transmission (*SORCS3, KCNQ5*). The function of *CHADL* remains to be fully elucidated and thus far it is only predicted to have a regulatory role in chondrocyte differentiation. Overall, presented results further indicate the role of these ten genes in GTS and other TDs, particularly of non-coding rare variants which make for the majority of the model.

The model was shown to effectively differentiate GTS patients from healthy members of large families and healthy samples from the same population. The best-fit model was based on the weighted overrepresentation of variants in these ten genes (AUC = 0.62, p-value = 0.02), whereas the next best model was the same with the only difference being the application of a MAF filter (MAF < 0.0001 AUC = 0.6). This indicated that the majority of important variants included in the model were rare with MAF below 0.01%. Indeed, the median MAF of all variants of the oligogenic risk model was 0.02. The majority of the variants included in the classifier are non-coding, mostly intronic. Although the interpretation of the role of non-coding variants is challenging and interactions at a distance cannot be excluded, it seems plausible that the identified variants affect the expression of nearby genes, most likely those included in the classifier (Zhang and Lupski 2015). The proposed risk model showed statistically significant sensitivity and specificity, albeit it was not perfectly accurate. This limitation may be explained by other variants contributing to the disease risk and the existence of external factors that influence the development of tics. Furthermore, we tested it on families where the closely related healthy individuals were expected to (and did) carry some of the risk variants. To investigate the genetic background of GTS in the families further, we also applied the common-variant PRS model based on GWAS data, which performed nearly at random (AUC = 0.47) at the same task as the oligogenic model.

When oligogenic scores were compared between groups, individuals diagnosed with non-GTS TDs had the mean value of the oligogenic risk score between healthy family members and their GTS-diagnosed relatives. There was a nominal statistically significant difference between the group of non-GTS TDs and their healthy relatives. This finding additionally validated the approach, as the non-GTS TDs group was not included in any steps of model development. Furthermore, this result is in concert with the idea that psychiatric disorders form a continuum of phenotypes rather than being discrete units and that GTS and other tic disorders constitute a spectrum of tic disorders with a common genetic background (Robertson et al. 2017; Taylor et al. 2019). This was further corroborated but the common-variant PRS where patients with severe GTS had the highest median value just like in the oligogenic score comparison. The term tic spectrum disorders was already suggested after a clinical study assessing symptoms of patients with GTS and chronic motor TD four that they existed on one spectrum of symptoms (Müller-Vahl, Sambrani, and Jakubovski 2019). The results of our study provide similar evidence but at the level of genomes.

This study has several limitations. One of the factors that may have influenced the model is the high genetic homogeneity of the Polish population (Jarczak et al. 2019). This could make the model population-specific. It is, therefore, necessary to confirm the results of this study with other patient populations. However, the discovery of increasing variant burden with the severity of symptoms should be generalizable regardless of the individual contributing variants. As unrelated controls we included a group of Polish elite sportsmen which was available locally to our team. Although this group was free of any neurological disorders it may not represent the overall Polish population. We aimed to minimize this limitation by adding groups of synthetic controls and using them in the feature selection step and including sportsmen only in the second step. Still, further WGS studies of substantially larger groups should provide an even better tool for oligogenic GTS risk prediction. Among GTS patients, 86% have other comorbid psychiatric disorders, mainly ADHD and OCD. They also exhibit emotional and behavioral problems, ASD, and cognitive deficits (Channon et al. 2006; Huisman-van Dijk et al. 2016; Robertson 2015; Verté et al. 2005). These comorbidities suggest that the underlying molecular, cellular, and neurophysiological etiology of GTS may be broadly applicable to a wide range of psychiatric disorders. We analyzed the oligogenic model in terms of being able to distinguish patients with and without comorbidities and concluded that the selected oligogenic model is not efficient in distinguishing any of the comorbidities apart from anxiety disorder suggesting it is likely specific for tic disorders. The association of variants in GTS risk genes and anxiety disorders should be further investigated in other cohorts.

Overall, the presented approach provides a promising path for further studies of the genomic basis of GTS. The obtained results support the concept that the additive effect of putatively deleterious variants in a small subset of key genes is a substantial risk factor of GTS and that TDs form a spectrum of diseases where increasing genetic risk burden is associated with more severe symptoms. We found that patients with GTS and severe tics that persisted into adulthood have the highest common and rare variant burden. We have validated results currently available in the literature and identified a range of rare non-coding variants not previously associated with GTS that could contribute to its etiopathology. The ability of the classifier to distinguish GTS-affected from healthy individuals within families is of particular importance, as patients with high genetic burden could be identified as being at increased risk of severe GTS persisting into adulthood. Although the current clinical utility of the presented model is limited, it provides an insight into the variant burden associated with typical as well as severe GTS.

## Supporting information

supplemental material description

## Declarations

### Ethics approval and consent to participate

The study was approved by the Ethics Committee of the Medical University of Warsaw (KB/2/2007, KB/53/A/2010, KB/63/A/2018) and The District Medical Chamber in Gdansk (KB-8/19) and has therefore been performed in accordance with the ethical standards laid down in the 1964 Declaration of Helsinki and its later amendments. All participants or their legal representatives gave written informed consent prior to inclusion in the study.

### Consent for publication

Not applicable

### Availability of data and materials

Datasets generated during the current study are available in the Supplementary Material. The main raw results consist of summary statistics of all genetic variants genotyped during Whole Genome Sequencing and are available in Supplementary Table S1. These summary statistics are provided as individual genotype-phenotype data are not publicly available due to the personal data protection act and the type of consent the study participants gave. All the code used in this project is available in the oligogenic model section in the projects’ <u>GitHub repository</u>.

## Authors’ contributions

PJ performed the clinical work and collected DNA samples from the patients; CZ, JPF, SG, and PC handled the patient samples and patient data processing; PC managed and provided access to DNA sample collection from local controls; SG and DH handled the sequencing and raw data collection; MP and DH performed WGS data preprocessing; MB performed WGS data analysis; MP, MK, and MP conceptualized the oligogenic model and interpreted its results; MP developed the polygenic tool; MZ calculated the PRS scores, MB prepared figures, tables and drafted the initial manuscript; CZ, JPF, MP, and PJ wrote sections of the manuscript; all authors reviewed the manuscript

## Data Availability

Datasets generated during the current study are available in the Supplementary Material. The main raw results consist of summary statistics of all genetic variants genotyped during Whole Genome Sequencing and are available in Supplementary Table S1. These summary statistics are provided as individual genotype-phenotype data are not publicly available due to the personal data protection act and the type of consent the study participants gave. All the code used in this project is available in the oligogenic model section in the project GitHub repository.

## Acknowledgements

We would like to acknowledge all the participants for their contribution to the study.

