## supplemental material description for "Oligogenic risk model for Gilles de la Tourette syndrome reveals a genetic continuum of tic disorders"

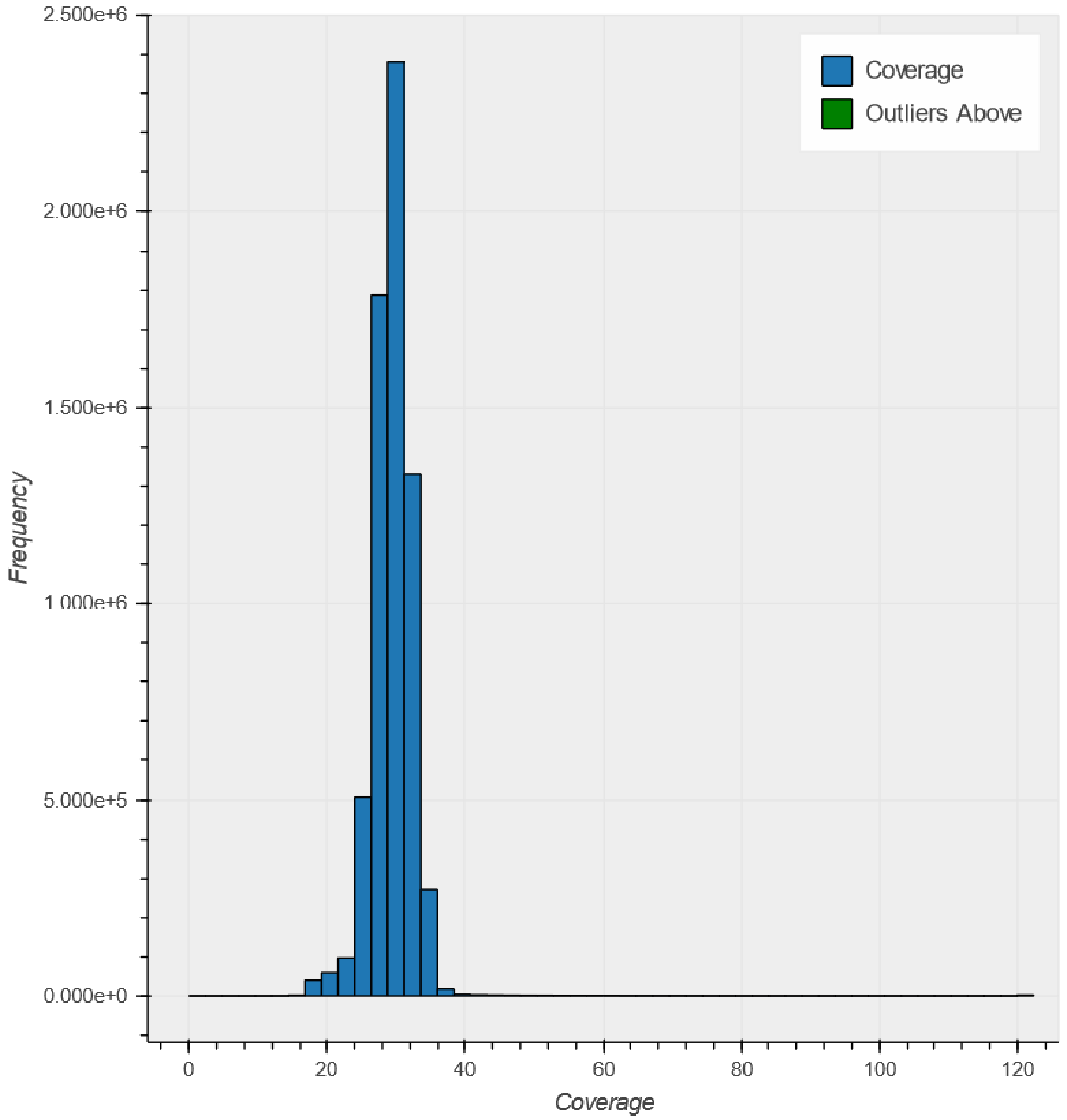


**Supplementary Figure S1.** Coverage histogram for WGS data used in this study (GTS and local controls groups, n = 278). Average coverage for the whole dataset was 29.7.

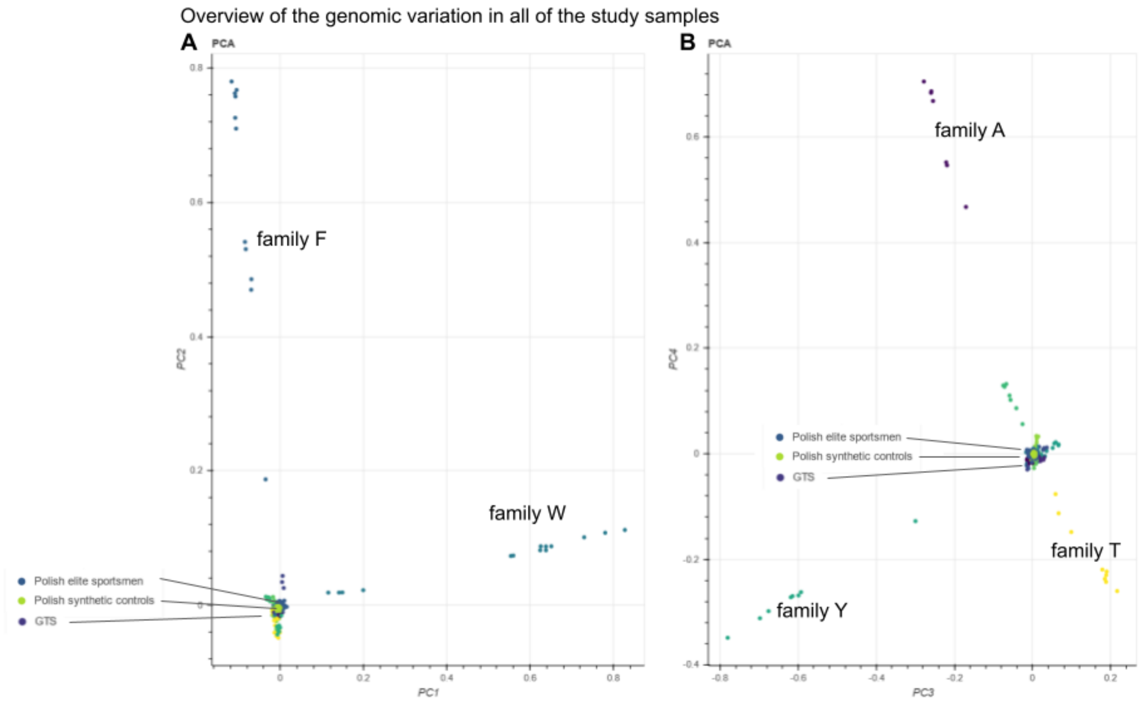


**Supplementary Figure S2.** PCA of all of the samples and synthetic controls included in the study, two plots of first 4 PCs are shown and indicate that kinship in the largest familites is the major factor determining the genetic variation in the cohort overall. **note: for the SKAT test PCs were recalculated for each synthetic control group and the test group of unrelated GTS patients**

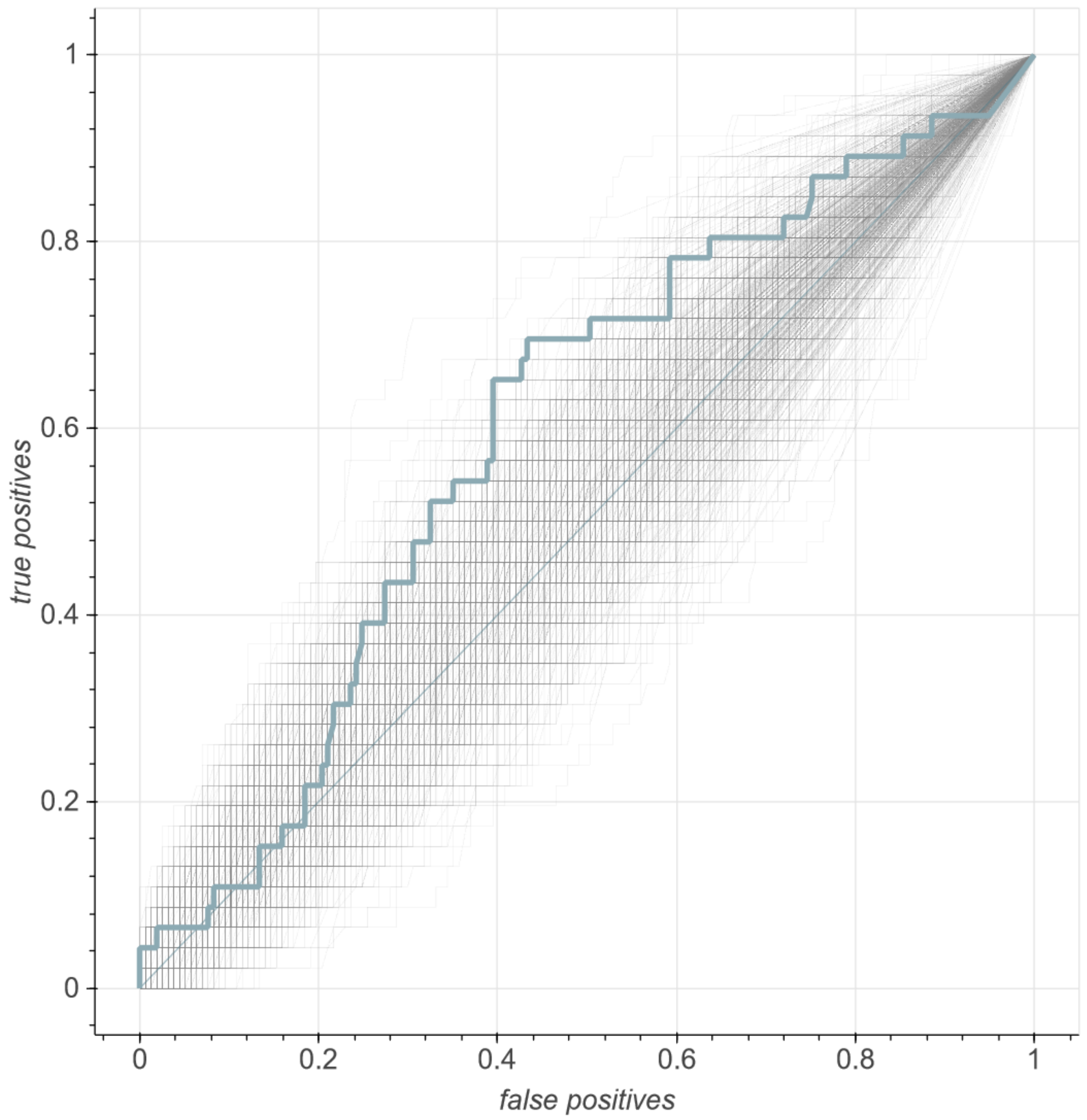


**Supplementary Figure S3.** Receiver-operator characteristics of the second best classifier, based only on very rare variants (AUC = 0.60) compared with 1000 classifiers based on randomly chosen gene lists. (p-value of the model based on the AUC = 0.04).

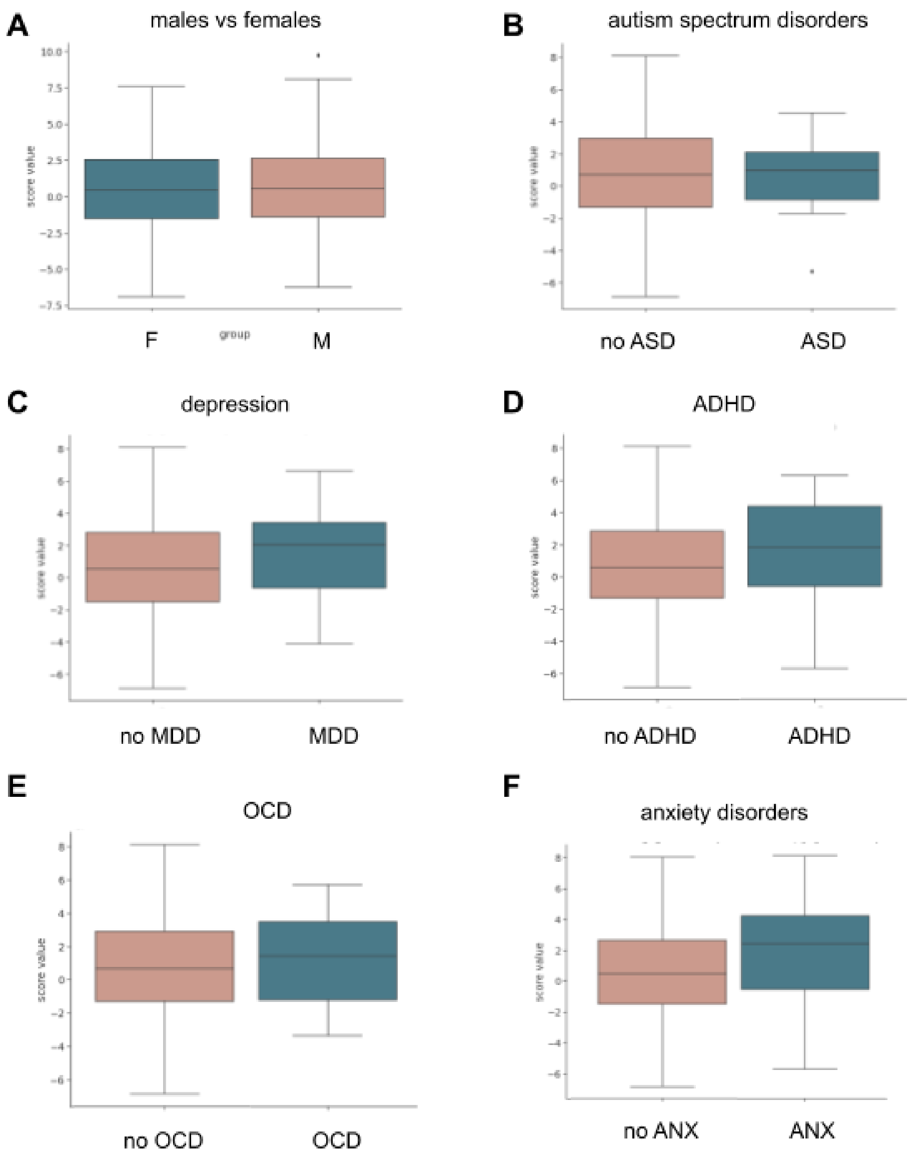


**Supplementary Figure S4. Oligogenic scores compared between sexes and different comorbidities.** Each group was compared with t-tests **A.** Oligogenic score per sex. T-test statistic = 0.13, p-value=0.89. **B.** ASD vs no-ASD: statistic=-0.33, p-value=0.74, **C.** Depression vs no depression: statistic=1.68, p-value=0.094, **D.** ADHD vs no ADHD: statistic=1.20, p-value=0.23, **E.** OCD vs no OCD: statistic=0.69, p-value=0.49, **F.** Anxiety vs no anxiety disorders statistic=2.20, p-value=0.03.

**Supplementary Table S1.** Summary statistics of genotype frequencies in each of the genotyped groups (healthy controls, polish simulated controls, familial cases (GTS and TDs) and sporadic cases (GTS) within/in the vicinity of 84 investigated candidate genes.

**Supplementary Table S2.** SKAT test results for the discovery group compared with 25 groups of synthetic controls. Size column - number of variants analyzed in the gene, other columns contain p-values for each of the 25 comparisons.

**Supplementary Table S3.** Full statistical results for Shapiro-Wilk, one-way ANOVA and pairwise T-tests for figure 3.

**Supplementary Table S4.** Assignment of the oligogenic model for each of the families included in the dataset. 'TRUE' - sample assigned as having a tic spectrum disorder, 'FALSE' - sample assigned as healthy.
